# A Survey of Provider Satisfaction of a New, Flexible Extended-Length Pharyngeal Airway to Relieve Upper Airway Obstruction During Deep Sedation

**DOI:** 10.1101/2020.11.20.20222018

**Authors:** Roxanne McMurray, Leah Gordan

**Affiliations:** Staff anesthetist, MNGI Digestive Care, Minneapolis, Minnesota, USA; Assistant Program Director, Nurse Anaesthesia Specialty; Clinical Assistant Professor at the University of Minnesota School of Nursing in Minneapolis, Minnesota, USA; Program Director, Nurse Anaesthesia Specialty; Clinical Assistant Professor at St. Mary’s University in Minneapolis, Minnesota, USA

**Keywords:** Airway management tools, airway complications, deep sedation, deep monitored anaesthesia care (MAC), obstructed airway

## Abstract

**Background:** Maintaining an open airway in a spontaneously-breathing patient under deep sedation, or deep monitored anaesthesia care, can be challenging. Specifically, current oral airways are not long enough to displace obstruction caused by redundant pharyngeal tissue, prompting external maneuvers by anesthetists that can impact patient outcomes and facility operational efficiency. As procedures increase at outpatient surgical centers, there is a need for an anesthesia provider-validated airway device that can sufficiently open an obstructed airway and maintain airway patency.

**Methods:** This prospective, multi-center user-experience survey evaluated anesthesia provider experience of a new airway device for adult patients with airway obstruction during deep sedation. The novel external airway has a longer flexible tubing allowing for displacement of pharyngeal tissue, smaller diameter to allow placement alongside an endoscopy bite block, and is manufactured with softer material to allow ease of insertion and patient comfort.

**Results:** Fifty-four anaesthetists at 15 hospital systems reported their experience of airway use in 86 cases. The novel airway device was 95% successful in establishing and maintaining a patent airway (n=68). Survey responses indicated that the airway was easy to place (93%), allowed for a “hands-off approach” (98%), and would improve airway management practice and patient outcomes (86%).

**Conclusions:** This pilot study demonstrated that the novel external airway is an effective and satisfactory method for anaesthesia providers to alleviate airway obstruction during deep sedation. Additional studies will be initiated to confirm efficacy and cost-effectiveness in patient populations and clinical environments that will most benefit from the new airway device.

## INTRODUCTION

Deep sedation, also referred to as deep monitored anaesthesia care (MAC), is an increasing anaesthesia modality in recent years for a type of cases (endoscopy, cardiovascular, orthopedics, podiatry, urology, GYN, etc.) designed to provide patient comfort and depression of consciousness during a procedure while preserving spontaneous ventilation(1, 2). Use of deep sedation is increasing due to several advantages it has over traditional general anaesthesia including decreased operating room time, faster patient recovery, decreased opioid use, reduced postoperative delirium, and less cardiac and pulmonary physiologic disruption (2-4). However, adverse events can occur, with respiratory events reported as the greatest cause of adverse outcomes during anaesthesia, precipitated by hypoventilation in more than a quarter of cases (5, 6). During upper endoscopies or colonoscopies under deep sedation, respiratory events due to inadequate oxygenation and/or ventilation were the leading cause of reported gastrointestinal procedure closed claims (7-9). In MAC endoscopy cases, airway-related complications related to hypoxemia can occur requiring airway maneuvers despite of preoxygenation (10). In addition, obese, sleep apneic, and elderly populations often undergo endoscopy procedures and these populations are at increased risk of upper airway obstruction because of reduced muscle tone and other upper airway complications during anaesthesia (9, 11-14).

If the airway is obstructed, anaesthesia professionals are trained to open and provide patency to improve airflow and oxygenation in patients’. There are a variety of patent airway devices utilized to maintain a patent airway, including nasopharyngeal airways (NPAs), NPA used orally, oropharyngeal airways (OPAs), supraglottic airway (SGA), and external noninvasive devices that maintain patient positioning. In addition, providers may consider performing physical positioning such as a chin lift or jaw thrust maneuver (15). Due to the risk of bleeding, NPAs are not ideal for anticoagulated patients. Current available OPAs are unable to stent open the airway beyond the tongue, allowing for collapse of soft redundant pharyngeal tissue despite the use of an airway device. As a result, anaesthesia providers often rely on chin lift or jaw thrust maneuvers throughout the procedure to maintain an open airway. This physical approach often results in patients reporting chin/jaw pain and bruising post-procedure, and providers reporting hand fatigue as well as limitation to tend to other critical tasks during MAC due to occupation of her/his hands (15). Furthermore, application of an external device, such as a continuous positive airway pressure (CPAP) or high-flow nasal oxygen can impact workflow as additional time and equipment may be required yet unavailable immediately. In an attempt to avoid such events altogether, workarounds to stent open the soft tissue have emerged such as oral placement of nasal airways due to convenience and longer-length tubing, which moves aside soft redundant pharyngeal tissue (15). Although these workaround approaches and off-label uses are intended to prevent adverse events associated with an obstructed airway, they may pose additional safety risks for the patient and potential liability issues for the health care provider and facility.

Workarounds have increased over the years because of changes in the human body and head size. Heads are larger and bodies are bigger, associating with a change in palate elongation and added pharyngeal and soft tissue (16, 17). The soft palate is considered a primary site of UAO in unconscious or anesthetized patients because it is the most compliant structure in normal and in obstructive sleep apnea (OSA) subjects (18-20). Existing oral airway equipment has not essentially been redeveloped over the last 100 years. The oral pharyngeal airway does not reach beyond the base of the tongue and the redundant pharyngeal soft tissue, leading to a deficit in providing a patent airway.

As the number of deep sedation/MAC cases continues to increase (2), an airway device that can quickly, efficiently, and effectively alleviate pharyngeal redundant tissue obstruction to improve ventilation and oxygenation may improve patient care, outcomes, and resource utilization. Given that the ease and speed of establishing upper airway patency is a major determinant of patient outcome, especially in the acute/emergency setting, provider satisfaction is a critical element of device appraisal. The aim of this preliminary evaluation was to introduce a new flexible extended-length pharyngeal airway device (16) in opening an upper airway obstruction during deep sedation to multiple centers and survey provider satisfaction from early adopters to establish proof-of-concept, collect initial user experience on human factors design, to inform subsequent provider training, and to inform design of a larger efficacy study.

## METHODS

This multi-center evaluator experience survey of early adoption was approved by a universal Institutional Review Board (IRB) for Human Subjects Research in March 2019. Anaesthesia providers who volunteered to trial this new airway device completed a survey tool to assess provider satisfaction of a new commercially-available upper airway device (McMurray Enhanced Airway (MEA); McMurray Medical, Minneapolis, MN).

The MEA is a novel upper airway device with numerous enhanced features relative to currently available airway management products (Figure 1). The MEA has longer flexible tubing allowing for displacement of pharyngeal tissue that oral airways are unable to reach, and avoiding the need for chin lift/jaw thrust maneuvers (16). The smaller diameter helps reduce stimulation and gagging and permits placement alongside an endoscopy bite block. The softer material, similar to that of a nasal airway, allows for ease of insertion and reduces potential oral injury associated with hard plastic oral airways (16). An elongated cushioned bite block is designed to prevent proximal airway collapse, allow flexibility of molar placement, and decrease the risk of dental damage (16). An optional connector can be connected to an anesthesia circuit or manual resuscitator, facilitating intraoral ventilation and aiding in situations such as difficult mask ventilation or when oxygen diffusion in the surgical field may present fire risk (16). Furthermore, the MEA was designed to reduce need for manual stationary maneuvers by providers such as chin lift or jaw thrust, thereby preventing potential provider-patient exposure of airborne droplets and increased staffing.

**Figure 1.**
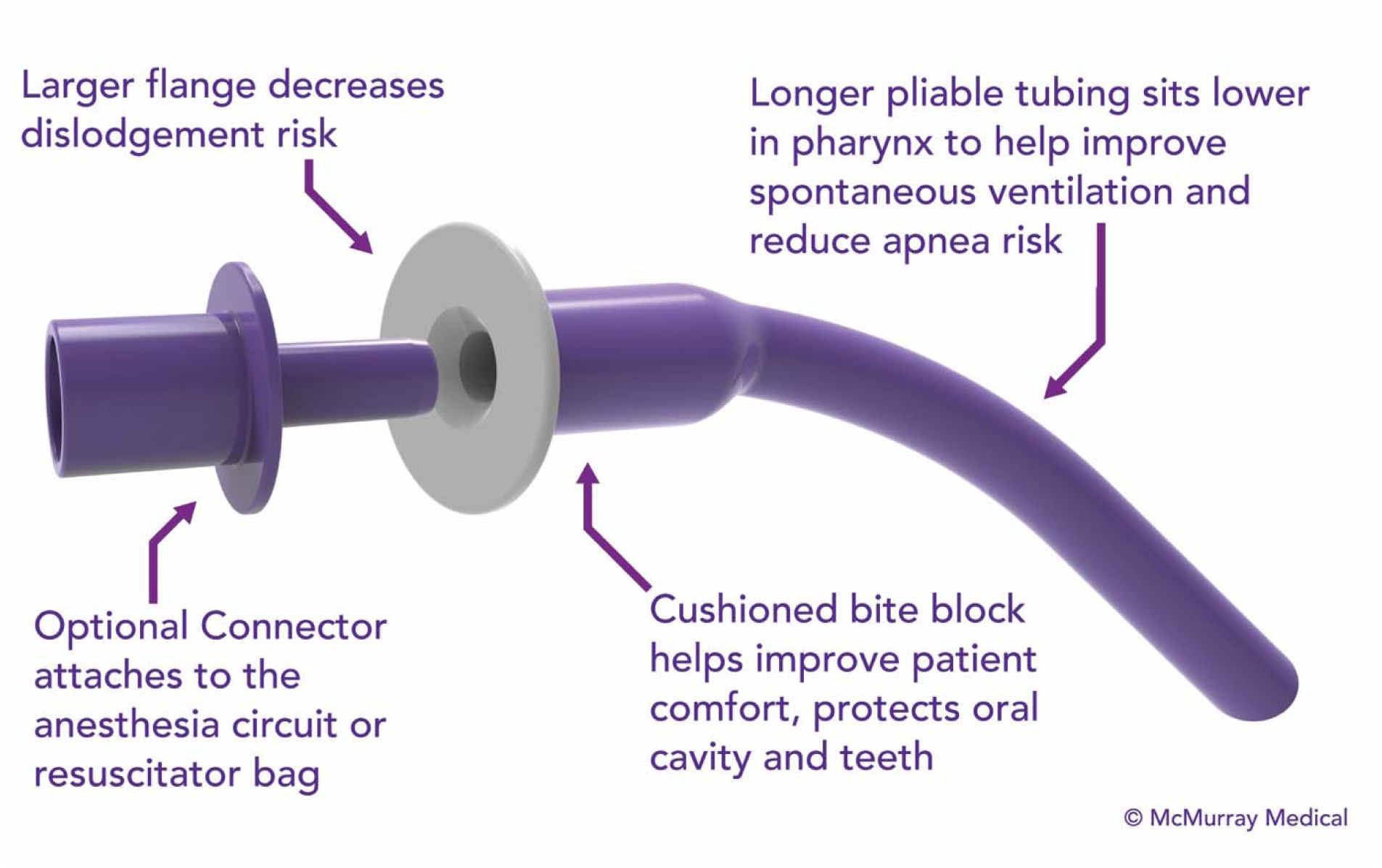
New flexible extended-length pharyngeal airway design (MEA). The MEA has longer flexible tubing allowing for displacement of pharyngeal tissue that oral airways are unable to reach. The smaller diameter helps reduce stimulation and gagging, and permits placement alongside an endoscopy bite block. The softer material allows for ease of insertion and reduces potential oral injury associated with hard plastic oral airways. An elongated cushioned bite block is designed to prevent proximal airway collapse, allow flexibility of molar placement, and decrease the risk of dental damage. An optional connector can be connected to an anesthesia circuit or manual resuscitator bag, facilitating intraoral ventilation and aiding in situations such as difficult mask ventilation or when oxygen diffusion in the surgical field may present fire risk (16).

Providers received device instructions for use (IFU) by training video and printed material to understand how to use and size the MEA. After using the MEA trial device in clinical practice, anaesthesia providers participated in completing the Use Survey Tool based on cases meeting the following criteria: inclusion criteria included adult patients (age >18) experiencing an obstructive airway under deep sedation; exclusion criteria included determining that the device size was inappropriate—too big or small—for the patient, since placing an improper size could be detrimental for the patient.

Surveys were distributed with the trial devices. As this was a pilot user feasibility study for the new extended pharyngeal airway device, recruitment of provider subjects was based on interest of using a novel airway device to benefit airway management when providing deep sedation in situations where upper airway obstruction became problematic. Surveys were voluntarily completed by the anaesthesia provider following each use. The initial survey (Phase 1) was developed to evaluate historic product usage for relief of airway obstruction, demographic details of patient experiencing airway obstruction leading to MEA use, anesthetic modality for MEA use (deep sedation or general anesthesia), and device satisfaction within each anesthetic modality with option for subjective free response. Based on Phase 1 response rate and provider feedback, Phase 2 surveys were limited to questions with ordinal responses that assessed device use satisfaction during deep sedation procedures only. One provider could complete multiple surveys to represent individual device performance and experience. Information related to patient, provider, and device placement frequency was not collected in order to expedite response rate to learn initial provider experience. Submitted surveys underwent response analysis by an independent statistician. Due to the nature of qualitative data, descriptive statistics were used to summarize study results. No objective measures of device clinical efficacy were collected.

## RESULTS

Fifty-four anesthesia providers from 15 different United States surgery locations (Phase 1 n=19, 5 different location; Phase 2 n=35, 10 different locations) voluntarily provided responses on device use and satisfaction (Phase 1 n=44; Phase 2 n=42). No adverse events reported.

### Phase 1

Nineteen providers voluntarily completed 44 device use surveys. (n=44 completed by 19 providers) indicated that all providers had experience with placement of traditional oral airway devices with 12 providers (63%) indicating experience with oral placement of nasal airways. Fifty percent of MEA placement occurred in patients with BMI >30. Patient demographics for MEA use are provided in Table 1.

**Table 1.**
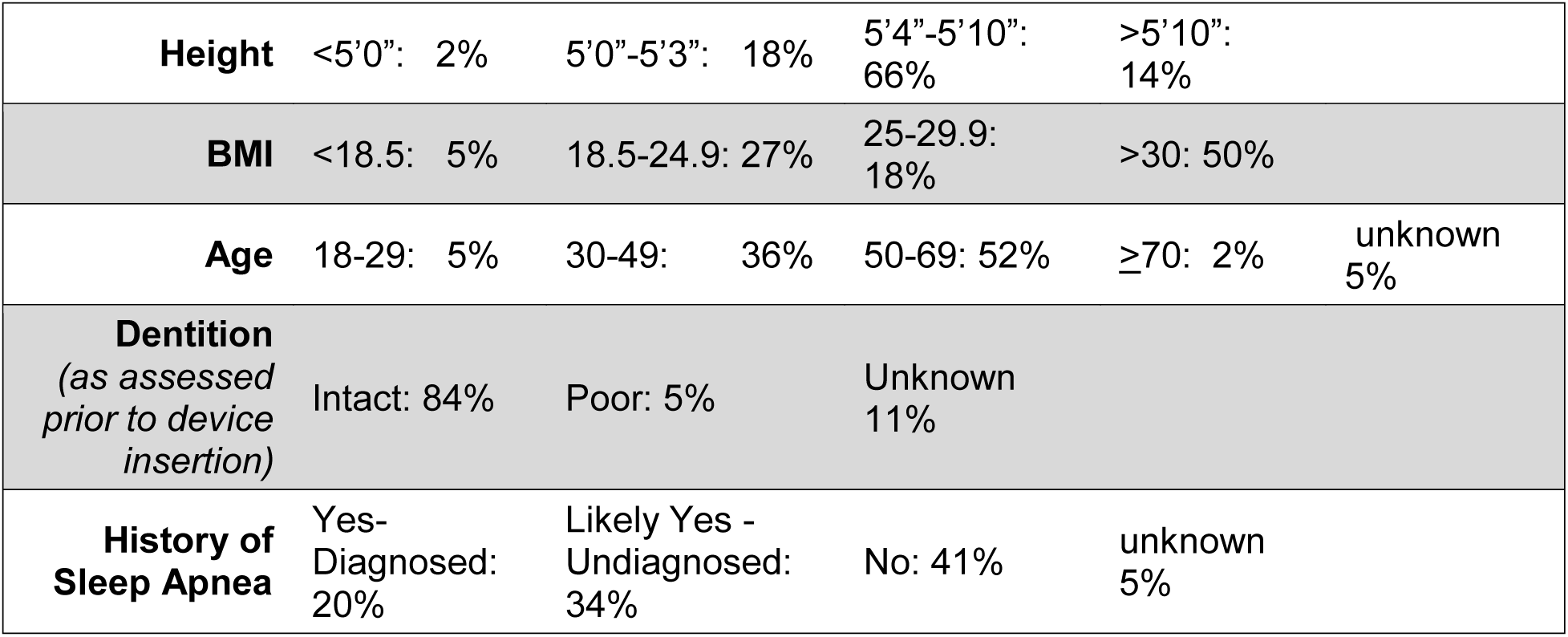
Phase 1 Responses on Patient Demographics for Device Use (n=44)

Out of the survey sections completed, 26 surveys indicated MEA use under deep sedation (59%) and included responses to the two questions related to device satisfaction during use in deep sedation conditions. Ninety-two percent (n=24) indicated that the MEA allowed a “hands off” approach and eliminated the need for chin/jaw lift maneuver. When asked if the MEA more readily maintains a patent airway during deep sedation compared to the airway currently used by that provider, 21 (81%) responses selected “Yes”, 2 responses selected “Somewhat” (7%), and 3 responses selected “No”(11%). Open responses related to “Somewhat” and “No” are shown in Table 2.

**Table 2.**
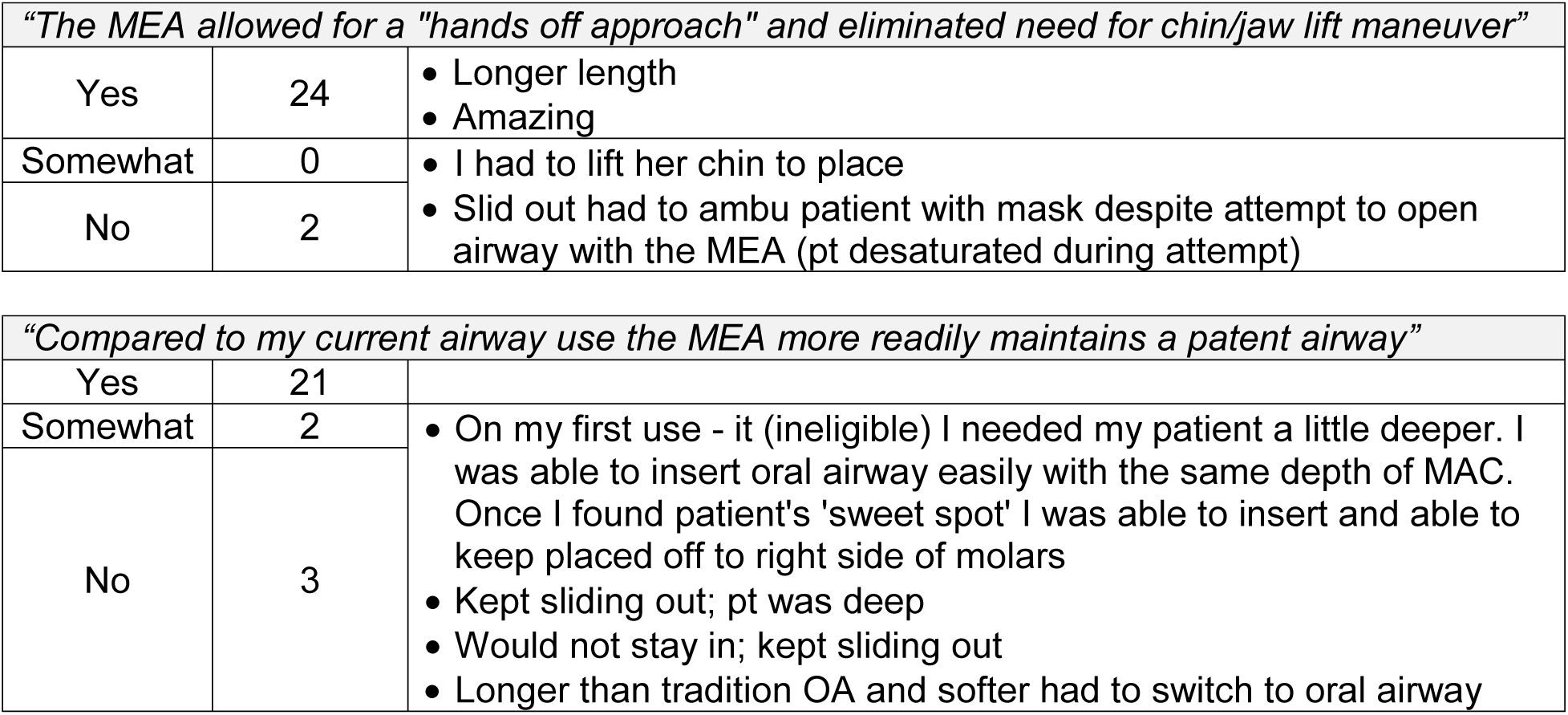
Phase 1 Device Satisfaction When Used During Deep Sedation (n=26)

Eighteen surveys indicated MEA use under general surgery (41%) to prevent collapsing of a Laryngeal Mask Airway (LMA) or Endotracheal Tube (ETT) upon extubation or to facilitate positive pressure intraoral ventilation. Two questions were asked related to device satisfaction during use in general anesthesia conditions. Response rate for general anesthesia questions was 66% (12/18) however 100% of respondents (n=12) indicated that the MEA prevented collapsing of the LMA or ETT. When asked if the MEA was easier to use compared to mask ventilation when facilitating positive pressure intraoral ventilation (n=6), 5 (83%) responses selected “Yes”, 1 response selected “No” (17%). Open responses related to “Yes” and “No” are shown in Table 3.

**Table 3.**
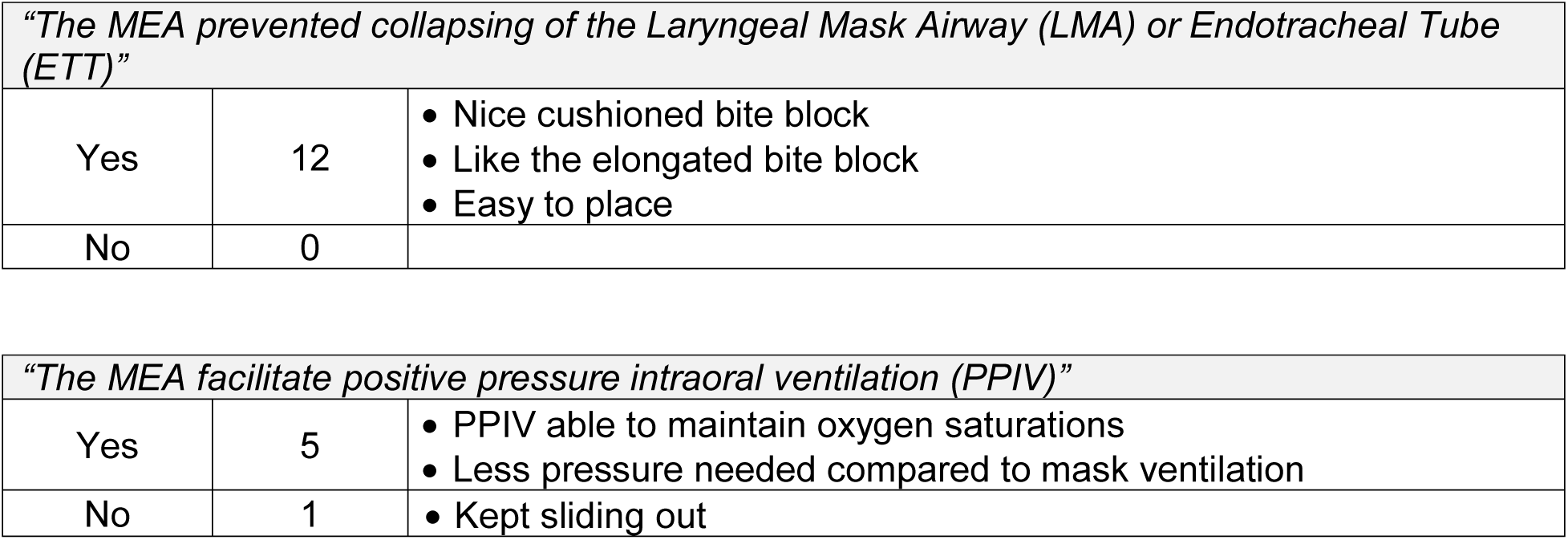
Phase 1 Device Satisfaction When Used During General Anesthesia (n=18)

| *"The MEA prevented collapsing of the Laryngeal Mask Airway (LMA) or Endotracheal Tube (ETT)"* |  |  |
| --- | --- | --- |
| Yes | 12 | • Nice cushioned bite block<br>• Like the elongated bite block<br>• Easy to place |
| No | 0 |  |

| *"The MEA facilitate positive pressure intraoral ventilation (PPIV)"* |  |  |
| --- | --- | --- |
| Yes | 5 | • PPIV able to maintain oxygen saturations<br>• Less pressure needed compared to mask ventilation |
| No | 1 | • Kept sliding out |

### Phase 2

Phase 1 responses were accompanied by responder criticism on barriers in voluntarily completing the patient demographics requested in Phase 1 survey. Thus, a Phase 2 survey was modified for subsequent device trials that evaluated device satisfaction during use in deep sedation only with 6 ordinal-response queries. Phase 2 surveys were completed for 42 device uses by 35 providers and results are presented in Table 4.

**Table 4.** Phase 2 Survey Responses on Device Performance and Satisfaction.

| *Phase 2 Survey Questions* | *Response Options* | *N* | *%* |
| --- | --- | --- | --- |
| **Was the MEA able to relieve airway obstruction?** | Yes | 42 | 100 |
|  | No | 0 | 0 |
| **Was the MEA used on an appropriate sized patient?** | Yes | 42 | 100 |
|  | No | 0 | 0 |
| **With the curve end facing the hard palate, how easy was it to place the MEA?** | Easy | 39 | 93 |
|  | Moderate | 3 | 7 |
|  | Difficult | 0 | 0 |
|  | Not Successful | 0 | 0 |
| **What position was the patient in? (circle all that apply)** | Supine | 31 | 74 |
|  | Lateral | 0 | 0 |
|  | Prone | 1 | 2 |
|  | Lithotomy | 1 | 2 |
|  | Trendelenburg | 0 | 0 |
|  | Reverse Trendelenburg | 0 | 0 |
|  | No response indicating patient position | 9 | 21 |
| **The MEA allowed for a “hands off approach” and eliminated need for chin/jaw lift maneuver.** | Yes | 41 | 98 |
|  | No | 1 | 2 |
|  | Not sure | 0 | 0 |
| **How satisfied are you with the MEA for deep MAC?** | Very satisfied | 38 | 90 |
|  | Somewhat satisfied | 0 | 0 |
|  | Satisfied | 4 | 10 |
|  | Dissatisfied | 0 | 0 |
|  | I did not use the MEA for deep MAC | 0 | 0 |
| **The MEA will improve my airway management practice.** | Yes | 36 | 86 |
|  | No | 0 | 0 |
|  | Not sure | 6 | 14 |

All phase 2 survey respondents (n=42; 100%) reported that the new airway device allowed for adequate ventilation and decreased upper airway obstruction. Out of the survey sections completed, 97% of responders (n=41) indicated that the device allowed for a “hands-off approach” and eliminated the need for chin lift or jaw thrust maneuvers. Thirty-nine responses (93%) selected that the device was easy to place and use under deep MAC, with 3 (7%) indicating it was moderately easy to place. Thirty-eight of Phase 2 responders (90%) surveyed were very satisfied with the new airway for deep MAC and the remaining four (10%) were satisfied, with no provider (0%) indicating dissatisfaction. When asked if the new device would improve airway management practice and patient outcomes, 36 providers who completed Phase 2 survey (86%) indicated Yes, while six (14%) said they were Unsure.

## DISCUSSION

In this preliminary evaluation of device use and user satisfaction, a new, flexible, extended-length pharyngeal airway device (MEA) to relieve obstructive airway under deep sedation was evaluated for usefulness and provider satisfaction. This is the first provider use evaluation for this innovative airway device and was initiated to gain preliminary insight on provider use and factors for adoption. Survey responses indicate that responding providers were satisfied with MEA’s performance in relieving upper airway obstruction during deep sedation.

As deep sedation administration increases inside and outside the hospital operating room (1, 2, 4, 17) risk for upper airway obstruction will also increase. Deep sedation administration can be challenging, leading to upper airway obstruction and its associated complications. Current airway practices have a void in this area, leading to an emergence of workarounds in deep sedation airway management (15). Improved airway management will contribute to better patient outcomes and patient satisfaction. Additionally, it may help reduce risk of litigation for providers and hospital systems, as inadequate ventilation and oxygenation are the source of more than one quarter of the reported MAC anaesthesia closed claims (6, 8).

There is a growing need for an airway device that can sufficiently open the upper airway obstruction and improve patient outcomes. Given that the ease and speed of establishing upper airway patency is a major determinant of patient outcome, provider satisfaction is a critical element of device appraisal and adoption into current workflow. Additionally, given the continued rise in prevalence of ambulatory anesthesia (specifically MAC), which presents with inherent time constraints, high patient turnover, and the necessity of timely and efficient discharge, the need for patent airway devices that can be applied efficiently and with diminished user error is crucial. Bhananker et al., performing a closed claims analysis of MAC, found that 41% (43/121) of claims were resultant of sub-standard care and could have been prevented by better provider performance (13). While provider performance is multifaceted, device user satisfaction is a central component. The assessment of provider performance and satisfaction has been established in studies evaluating airway devices included assessment of provider performance and/or satisfaction (18-22) compared conventional mask and OPA ventilation to that of an esophageal obturator airway device in patients (n=10) who were scheduled for surgery on an extremity, inguinal herniorrhaphy, or cystoscopic examination under general anesthesia and included ordinal ranking of operator effort as reported by the provider and an observer. Dob et al. (1999) compared the efficacy of a laryngeal mask airway (LMA) (n=26) to that of a traditional OPA device (n=26) in maintaining airway patency following tracheal extubation (middle ear surgery) and assessed provider reporting of ease of airway maintenance, or ‘work required’, via an ordinal scale (19). Similarly, Khosravan et al. also compared the efficacy of an LMA (n=18) to that of a traditional OPA airway (n=17) in managing failed intubation in the pre-hospital emergency setting, reporting time spent, number of attempts, and the need for head positioning for each respective device (20). Shaikh et al. (2016) performed a study examining provider success in placing either a traditional OPA device or an oropharyngeal device possessing circumferential oropharyngeal seal cuff airway device in patients (n=60) who were scheduled for surgery under general anesthesia, with ease of airway insertion was reported on a 10-point ordinal scale by the provider (21). Xiao et al. (2016) evaluated the efficacy and safety of an NPA device (n=130) to that of a nasal oxygen tube (n=130) in obese patients receiving general anesthesia and undergoing painless gastroscopy with physician and anesthetist satisfaction of the collective anesthesia protocol reported on a 10-point ordinal scale (22). Recently, De Rosa et al. (2019) included subjective assessment of device insertion when evaluating an LMA with a tracheal cannula in general anesthesia conditions (23).

This pilot evaluator survey of the MEA contributes to the reporting of anaesthesia provider satisfaction during evaluation of airway device use. Specifically, these preliminary surveys indicate that MEA allowed adequate ventilation and decreased upper airway obstruction, enabled an approach that allowed minimal manual stability (“hands-off”) by the provider. Furthermore, the majority of airway experts who participated in this study and provided survey responses had satisfactory experiences with this device.

Limitations to this pilot study include the number of participants, convenience sampling with trial airway devices, subjective assessments, and low response rate during initial surveying, confirming previous discussion on complexity of gaining user experience of nursing innovations (24-26). Thus, the responses that were collected may represent a selection bias and not be reflective of every provider’s experience with the MEA. Additionally, due to the intent to quickly collect initial feedback on provider experience to better understand airway device utilization, the number of patients attempted for MEA use was not recorded. Furthermore, patient-specific procedure details were not requested to identify opportunities for clinical performance improvement. However, these initial documents of device use and provider opinions provide rationale to continue product development and assessment.

Future robust studies will further assess the utility of the new pharyngeal airway in controlled studies conducted by independent principal investigators with subjective and objective data collection on provider experience. Although this survey focused on the device’s ability to improve ventilation under deep sedation, more research is warranted to determine the usefulness in other airway management situations such as post-extubation or in patients with compromised airways outside the procedure and operating room and working efficiently with other medical devices such as fitting alongside an endoscopy bite block, connecting to anaesthesia circuits or manual resuscitators (27). Additionally, future studies will aim to compare the efficacy of the new device to other currently available airway devices in prospective, randomized, controlled studies.

## Supporting information

Universal Institutional Review Board

## Data Availability

All the data is available upon request.

https://b19e721b-fcc9-43eb-b742-beec9e9153b4.filesusr.com/ugd/42fff9_d59d793e392e45c69cc0035b33b87588.pdf

## ACKNOWLEDGMENTS

The authors would like to thank the providers who volunteered to participate in the surveys and Amy Moore and Jana Stender for their contributions during the manuscript review process. No funding was necessary for the implementation of the project.

## COMPETING INTERESTS

Roxanne McMurray is the inventor of the McMurray Enhanced Airway (MEA) and Vice President of Clinical Management at McMurray Medical.

## Notes

### Clinical Trial

NCT04419883

### Author Declarations

This study was approved by a Western Institutional Review Board (IRB) for Human Subjects Research in March 2019.

