## Supplementary material for "A Survey of Provider Satisfaction of a New, Flexible Extended-Length Pharyngeal Airway to Relieve Upper Airway Obstruction During Deep Sedation": Universal Institutional Review Board

WIRB
Global Leader | Proven Expert

March 4, 2019

Roxanne McMurray, DNP
McMurray Medical, LLC
804 Amble Road
Shoreview, MN  55126

Dear Dr. McMurray:

SUBJECT: IRB EXEMPTION— HIPAA FULL WAIVER OF AUTHORIZATION AND REGULATORY OPINION
McMurray Medical, LLC
Sponsor Protocol #: MEA-001-01
Protocol Title: McMurray Enhanced Airway (MEA) Product Evaluation Survey

On, March 3, 2019 Western Institutional Review Board (WIRB) **approved** a request for a waiver of authorization for use and disclosure of protected health information (PHI) for the above-referenced research. This review was conducted through expedited review.

**WAIVER OF HIPAA AUTHORIZATION**

WIRB determined that documentation received from you satisfies the three requirements for a waiver of authorization. These requirements are:

1. The use or disclosure of the PHI involves no more than minimal risk to the individuals, based on the following elements:

   a. An adequate plan to protect identifiers from improper use and disclosure;
   b. An adequate plan to destroy the identifiers at the earliest opportunity consistent with conduct of the research (unless there is a health or research justification for retaining the identifiers, or such retention is otherwise required by law); and
   c. Adequate written assurances that the PHI will not be reused or redisclosed to any other person or entity, except as required by law, for authorized oversight of the research project, or for other research for which the use or disclosure of PHI would be permitted by HIPAA.

2. The research could not be practicably conducted without access to and use of the PHI; and

3. The research could not practicably be conducted without the waiver.

The Board has determined that this waiver of authorization for the use and access of the protected health information as described in the above referenced protocol, and in the information provided in the submitted waiver of authorization form, is necessary for conduct of this research.

**REVIEW OF EXEMPTION REQUEST**

This is in response to your request for an exempt status determination for the above-referenced protocol. Western Institutional Review Board's (WIRB's) IRB Affairs Department reviewed the study under the Common Rule and applicable guidance.

We believe the study is exempt under 45 CFR § 46.104(d)(2), because this research involves a survey of CRNAs and Anesthesiologists on the use of products for disposable airway device intended for single use only. No information is being submitted to the FDA. This study will not collect identifiable participant information.

This exemption determination can apply to multiple sites, but it does not apply to any institution that has an institutional policy of requiring an entity other than WIRB (such as an internal IRB) to make exemption determinations. WIRB cannot provide an exemption that overrides the jurisdiction of a local IRB or other institutional mechanism for determining exemptions. You are responsible for ensuring that each site to which this exemption applies can and will accept WIRB's exemption decision.

Please note that any future changes to the project may affect its exempt status, and you may want to contact WIRB about the effect these changes may have on the exemption status before implementing them. WIRB does not impose an expiration date on its IRB exemption determinations.

If you have any questions, or if we can be of further assistance, please contact Tanna M. MacReynold, C.I.P., at 360-252-2869, or.

Sincerely,

[signature] FOR

Kelly FitzGerald, PhD
Vice President, IRB Affairs

TMM:jca
D2 and Hipaa-Exemption-McMurray (03-04-2019)
cc: Cassandra Latimer, MS, Latimer Consulting LLC
WIRB Accounting
WIRB Work Order #1-1162336-1

This document electronically reviewed and approved by MacDonald, Currien on 3/4/2019 5:10:37 PM PST. For more information call Client Services at 1-360-252-2500.
